# Yellow fever in Ghana: Predicting emergence and ecology from historical outbreaks

**DOI:** 10.1101/2024.01.29.24301911

**Authors:** Seth D. Judson, Ernest Kenu, Trevon Fuller, Franklin Asiedu-Bekoe, Alberta Biritwum-Nyarko, Lee F. Schroeder, David W. Dowdy

## Abstract

Understanding the epidemiology and ecology of yellow fever in endemic regions is critical for preventing future outbreaks. Ghana is a high-risk country for yellow fever. In this study we estimate the epidemiology, ecological cycles, and areas at risk for yellow fever in Ghana based on historical outbreaks. We identify 2371 cases and 887 deaths (case fatality rate 37.4%) from yellow fever reported in Ghana from 1910 to 2022. Since implementation of routine childhood vaccination in 1992, the estimated mean annual number of cases decreased by 81% and the geographic distribution of yellow fever cases also changed. While there have been multiple large historical outbreaks of yellow fever in Ghana from the urban cycle, recent outbreaks have originated among unvaccinated nomadic groups in rural areas with the sylvatic/savanna cycles. Using machine learning and an ecological niche modeling framework, we predict areas in Ghana that are similar to where prior yellow fever outbreaks have originated based on temperature, precipitation, landcover, elevation, and human population density. We find differences in predictions depending on the ecological cycles of outbreaks. Ultimately, these findings and methods could be used to inform further subnational risk assessments for yellow fever in Ghana and other high-risk countries.

**Author Summary:** Yellow fever is a viral hemorrhagic fever transmitted by mosquitoes in Africa and South America through different ecological transmission cycles. While West Africa has had the most cases of yellow fever, less is known about the epidemiology and ecology of yellow fever among countries in this region. Ghana has had multiple yellow fever outbreaks, including a recent outbreak in 2021-2022. In this study we estimate cases and deaths due to yellow fever in Ghana, compare the ecological cycles of outbreaks, and predict future areas at risk based on prior yellow fever cases and environmental conditions. We find that the populations at risk for yellow fever in Ghana have changed over the past century and that different ecological factors influence the risk of future emergence. Understanding these changes and the nuances of yellow fever epidemiology and ecology within countries will be important for future outbreak preparedness.

## Introduction

Yellow fever (YF) outbreaks continue to occur in Africa and South America despite an effective vaccine having been created nearly a century ago. Yellow fever is a viral hemorrhagic fever caused by yellow fever virus (YFV), a mosquito-borne flavivirus. In Africa, YFV circulates through three transmission cycles. In the sylvatic cycle, forest-dwelling *Aedes* spp. mosquitoes infect non-human primates (NHPs) and intermittently transmit YFV to humans after feeding on infected NHPs [1]. In the savanna or intermediate cycle, humans are infected by mosquitoes in forest border areas, and there is human-to-human and NHP-to-human transmission via mosquito vectors [1]. In the urban cycle, there is only transmission among humans from the anthropophilic mosquito *Aedes aegypti*, which breeds in water-containing vessels in urban sites [1]. While all three transmission cycles cause human cases of YF in Africa, it remains unknown how each cycle contributes to the burden of YF, limiting targeted interventions and predictive models [2,3].

Among infectious diseases in the WHO Africa Region (AFRO), YF caused the fifth most outbreaks in 2020 [4]. And while there are more cases of YF in West Africa compared to anywhere else in the world, a recent systematic review found only 12 studies of YF incidence and mortality within African countries [5]. Compounding the limited knowledge of the historical burden of YF, there are gaps in predicting future emergence of YF within African countries, and it is unknown how each of the three transmission cycles (sylvatic, savanna, and urban) contribute to YF outbreaks.

There is a contrasting situation in South America, where all outbreaks of YF in Brazil have been attributed to the sylvatic cycle since 1942 and there are known seasonal and climatic drivers of outbreaks [6]. In addition, some species of New World NHPs develop lethal infections due to YFV, and the finding of moribund or dead NHPs can indicate an epizootic outbreak, signaling impending sylvatic YF outbreaks among humans [1]. In contrast, most YFV infections among Old World NHPs in Africa are believed to be inapparent or subclinical, and therefore outbreaks from sylvatic and savanna transmission occur without alerting indicators in Africa. Initial sylvatic or savanna outbreaks could lead to epidemics if the urban cycle subsequently becomes established. Therefore, understanding the historical epidemiology and ecology of YF is crucial to predicting future emergence among YF-endemic African nations.

There is also concern for YF outbreaks within African countries to cause epidemics. In 2016 a YF outbreak in Angola spread to neighboring countries, and the epidemic response required more than 28 million YF vaccines, exhausting the global YF vaccine supply [7]. In response to this YF epidemic, the WHO, GAVI, and UNICEF developed the Eliminate Yellow Fever Epidemics (EYE) strategy with the aim of eliminating yellow fever epidemics by 2026 [7]. The core objectives of the EYE strategy are to (1) protect at-risk populations, (2) prevent international spread, and (3) contain outbreaks rapidly [8]. Central to achieving these objectives is understanding the regions and populations at greatest risk for YF outbreaks. Ghana is among the 27 countries identified by EYE as being high-risk for YF and has had multiple YF outbreaks.

Understanding the contexts of YF outbreaks in Ghana and identifying areas at future risk could inform resource allocation for vaccination campaigns, vector control, and diagnostic testing. Modeling tools could aid risk assessment for YF by predicting areas at risk based on prior outbreaks. For zoonotic and vector-borne diseases like YF, the locations of cases, vectors, or hosts can be used to create spatial models which can be translated into risk maps [9]. For example, researchers have predicted the local risk for sylvatic YF in Brazil from models based on human and NHP cases as well as ecological factors [10,11].

Through combining epidemiological data from historical YF outbreaks with local ecological knowledge, we aim to predict the habitats and factors associated with YF in Ghana. Because there are limited data on the multiple *Aedes* spp. and NHPs that may propagate YFV circulation in Ghana, we use a machine learning approach incorporating confirmed human YF cases as inputs and multiple abiotic and biotic covariates as explanatory variables. Ultimately, our overall goals are to (1) review the historical epidemiology of YF outbreaks in Ghana, (2) assess contributions of different YF ecological cycles to these outbreaks, and (3) identify areas at risk for YF emergence based on this historical and ecological understanding.

## Methods

### Setting

The Republic of Ghana is a country in West Africa on the Gulf of Guinea, bordered by Côte d’Ivoire, Burkina Faso, and Togo. Ghana consisted of 10 regions until December 2018 when the Brong-Ahafo, Northern, Volta, and Western regions were split to make a total of 16 regions. For the purposes of this study, we used the current geographic boundaries of the 16 regions and 260 districts in Ghana. For outbreaks prior to 2019 with available geographic information, we re-classified these locations to align with the current terminology for regions and districts. We also compared outbreak locations by the five recently proposed agro-climatic zones for Ghana, which are named from north to south: Sudan Savannah, Guinea Savannah, Transition Zone, Forest, and Coastal [12]. These zones encompass geographic regions with similar temperature and precipitation patterns [12].

The Government of Ghana introduced the YF vaccine as part of the Expanded Program on Immunization (EPI) in 1992, making it a routine immunization for children in Ghana. Although the WHO estimates that 95% of the population in Ghana has been vaccinated against YF [13], Ghana had a recent YF outbreak from October 2021 to February 2022 with 70 confirmed cases and 35 deaths [14]. The origin and epicenter of the outbreak was in the West Gonja district of the Savannah region, among unvaccinated nomadic populations, from which the outbreak spread to other regions [15,16]. We reviewed the epidemiology of this latest outbreak and prior outbreaks using the methods described below.

### Identifying YF Outbreaks in Ghana

To identify YF outbreaks in Ghana and the locations of corresponding cases, we searched PubMed, EBSCO, Google Scholar, and Scopus for literature using the search terms “yellow fever” AND “Ghana.” We also searched the ProMed archives using the same search terms. To find WHO YF reports, we used the WHO’s Institutional Repository for Information Sharing (IRIS) and queried the Medical Subject Heading (MeSH) “yellow fever”, which included the Disease Outbreak News (DON) and Weekly Epidemiology Record (WER). This also included a 2000 WHO report which included YF case reporting among countries from 1950-1998 [17]. We also reviewed additional WHO documents, including a 2005 report on YF in Ghana [18] which was referenced in a review of arboviruses in Ghana [19]. Finally, we reviewed publicly available online reports from the Ghana Health Service (GHS) and the Ghana Weekly Epidemiological Report. The literature search was conducted up to October 20th 2023. We collated all of the data from the aforementioned sources into a dataset of reported annual YF cases, deaths, and case fatality rates (CFR) for individuals in Ghana, as well as a dataset of outbreaks in Ghana (locations, month of outbreak onset, and reactive vaccination campaigns) [20].

### Estimating Annual YF Cases and Outbreak Characteristics

For the period from 1910-1950, we extracted YF cases, deaths, and outbreak characteristics primarily from a single source (Scott 1965) [21]. For 1950-2022, we compared YF cases and deaths between multiple sources, including the primary literature and WHO reports. If there was a discrepancy in annual cases/deaths, we used the total cases/deaths reported in the primary literature, given delays between reporting cases and confirmatory testing.

We excluded cases from 2003 which were reported only in the WHO 2005 report [18] because we could not find these cases referenced in another source. We were also unable to find deaths reported for 31 cases in 2012 [22]. If the source distinguished between reported and confirmed cases, as well as diagnostics used, we also included this information in our dataset. For our calculations of disease burden and CFR, we used overall reported cases and deaths.

### Categorizing YF Outbreaks According to Likely Transmission Cycles

For outbreaks with sufficient epidemiological information (including location details, diagnostic method, and vector surveillance), we categorized the outbreaks as likely occurring due to the urban, sylvatic, or savanna cycles [20]. If vector surveillance identified sufficient *A. aegypti* based on the indices described below, or if the outbreak was concentrated around urban dwellings or structures (i.e. wells or water storage containers) then these outbreaks were attributed to the urban cycle. Outbreaks where vector surveillance revealed insufficient *A. aegypti* and/or cases originated in forest or savanna habitat were attributed to the sylvatic or savanna cycle. Given limitations in distinguishing between the sylvatic and savanna cycles, these were considered together for our modeling analyses.

For categorizing urban YF, we considered the following vector indices: (1) house index (% of houses with at least one positive breeding place), (2) container index (% containers with *A. aegypti* larvae), and (3) Breteau index (# of positive larval breeding places per 100 houses) [23]. There is a high risk for urban YF when the house index > 35, container index > 20, and Breteau index > 50 [23]. If the Breteau index is between 5 and 50, then there is considered to be sufficient *A. aegypti* to cause an outbreak. There is unlikely to be urban transmission of YFV when the house index <4, container index <3, and Breteau index <5 [23].

### Georeferencing YF Occurrences

To obtain occurrence data for our models, we identified the locations of laboratory confirmed human YF cases in Ghana. A panel of YF experts had previously determined that one of the most important risk factors for YF in the African region was confirmed YF cases since 1960 [24]. Therefore, we included only confirmed cases since 1960 for our models. We georeferenced each case to the town of residence; if this was unknown then we used the location of the healthcare facility where the patient was evaluated. To improve precision, we excluded cases that were only referenced at the region or district level. Our methods for georeferencing are further described in S1 text.

To mitigate sampling bias, we treated multiple cases at the same location in a year as a single occurrence. We also excluded locations that where within 1 km of another occurrence, given our spatial resolution of covariates at 30 arc seconds (∼1 km^2^). We categorized the transmission cycles of occurrences as described above.

Through this process we identified 21 occurrences for YF in Ghana from 1960-2022. We categorized six of these occurrences as being due to the urban cycle given positive vector indices for *A. aegypti*. Additional details on the occurrences are available in the online dataset and S1 text. To compare the regional distribution of YF cases as well as districts where YF outbreaks have originated, we created maps in ArcGIS Pro (https://www.esri.com).

### Modeling Habitat Suitability for YF Emergence

To identify suitable habitats for YF emergence, we built on a previously described approach that is optimized for small sample sizes to identify habitat suitability for pathogen spillover [25]. We also used a similar methodology and covariates as those that were recently used to model YF emergence in Brazil [11]. We used Maximum Entropy Species Distribution Modeling (Maxent) version 3.3.3 [26], a machine learning algorithm commonly used in species distribution modeling and ecological niche modeling, to predict the distribution of a species based on presence locations as input and multiple ecological covariates. We chose Maxent given that it has outperformed other modeling algorithms for small sample sizes with presence-only data, including in environments in Africa [27]. Our aim was to model the environments suitable for YF emergence in Ghana, as well as to explore the relative importance of different abiotic, biotic, and human covariates. Therefore, instead of making the stringent assumptions to interpret Maxent output as an ecological niche, we instead built our models to indicate habitat suitability [28].

We modeled YF habitat suitability as a function of multiple covariates (Table 1). These covariates were identified as factors that could influence the ecological cycles of YF and included (1) abiotic covariates: 19 bioclimatic variables and elevation (WorldClim 2.1, https://www.worldclim.org) [29], (2) biotic covariates: landcover (MODIS MCD12Q1, https://www.earthdata.nasa.gov) [30] and NHP species richness (https://sedac.ciesin.columbia.edu/data/set/species-global-mammal-richness-2015) [31], and (3) human population density (https://sedac.ciesin.columbia.edu/data/set/gpw-v4-population-density-adjusted-to-2015-unwpp-country-totals-rev11) [32].

**Table 1.** Explanatory covariates for YF habitat suitability models.

| Covariate | Details | Resolution | Source |
| --- | --- | --- | --- |
| 19 bioclimatic variables | Gridded climate data 1970-2000 | 30 arc seconds/1 km<sup>2</sup> | Worldclim 2.1 |
| Elevation | Derived from the Shuttle Radar Tomography (SRTM) elevation data | 30 arc seconds/1 km<sup>2</sup> | Worldclim 2.1 |
| Species richness of Old World monkeys (Cercopithecidae) | Gridded values of the number of species of Cercopithecidae from the IUCN Red list | 30 arc seconds/1 km<sup>2</sup> | Global Mammal Richness Grids, 2015 Release (2013) (SEDAC) |
| Landcover | Moderate Resolution Imaging Spectroradiometer (MODIS) Land Cover from 2021-2023 | 500 m resolution re-scaled to 1 km | MODIS MCD12Q1 NASA |
| Human population density | 2020 UN adjusted population density | 30 arc seconds/1 km<sup>2</sup> | UN WPP-Adjusted population density v4.11 (SEDAC) |

To identify potential NHPs that could propagate the sylvatic/savanna cycles, we referenced the IUCN Red List (https://www.iucnredlist.org/) and identified 17 species of NHPs in Ghana. Of these species, six had epidemiological and laboratory evidence of YF infection at the species level (*Cercopithecus mona*, *Colobus vellerosus*, *Erythrocebus patas, Pan troglodytes*, *Papio anubis*, *Perodicticus potto*), and three had evidence at the genus level (*Cercopithecus lowei*, *Cercopithecus petaurista*, and *Cercopithecus roloway*) [24]. Eight of these nine species are members of the Cercopithecidae family. The exception was *P. troglodytes*, which is not a common species in Ghana and is unlikely to have a significant role in YF ecology [24]. Therefore, we evaluated species richness of Old World monkeys (Cercopithecidae) as a covariate. Further details about the covariates are available in S1 text.

We created one set of models with all YF occurrences undifferentiated by ecological cycle, to predict areas with habitat suitability for YF irrespective of cycle. We created another set of models which excluded the six urban occurrences that had confirmed positive *A. aegypti* vector indices. For our model parameters, we used a leave-one out cross-validation (LOOCV) method that is optimized for small sample sizes [25,33]. To make our models comparable to recent Maxent models for YF from Brazil, we used similar Maxent parameters and methods for selecting covariates [11].

We ran our initial models in Maxent with all covariates, using a jackknife procedure to evaluate relative covariate importance. For our initial models, we used the default Maxent settings and additionally selected random seed, replicated run type as cross validate, replicates equal to the sample size, and set the maximum iterations to 100,000. To identify collinearity among covariates, we used a correlation matrix in ArcGIS Pro. We removed covariates that had >70% correlation as well as those with less than 5% percent contribution and permutation importance. We used both of these metrics and the jackknife test to compare the relative importance of covariates. Percent contribution was deemed less informative than the other metrics since it is determined heuristically and depends on the sequence of covariates [34].

Following this exclusion of potentially less-relevant covariates, we re-ran the models using the same LOOCV approach and settings. We used the cumulative output of the Maxent models as a representation of habitat suitability, where the value of each cell is the sum of all raw values less than or equal to the value for that location and rescaled from 0 to 100 [28]. The cumulative output can be used to create binary maps and determine omission rates for the models. For the binary maps and omission rates we used the minimum training presence cumulative threshold, which corresponds to the minimum habitat suitability in the training data. To compare the relative performance of the Maxent models we used the area under the receiver operating characteristic curve (AUC), which ranges from 0 to 1 and reflects the probability that randomly chosen presence locations are ranked higher than background points [28].

## Results

We identified 2371 cases and 887 deaths due to YF in Ghana from 1910 to 2022. The overall mean CFR was 37.4%. The first definitive outbreak of YF in Ghana occurred in 1910. From 1910-1960 there were 569 cases and 335 deaths (CFR 58.9%). In 1945 YF vaccination became required for foreigners in Ghana, and the first reactive mass vaccination campaign for YF in Ghana occurred in 1951. We identified 12 reactive vaccination campaigns from 1951-2022 (Fig S1).

We successfully identified the regions in which 1722/1802 (95.6%) of reported cases from 1960 to 2022 occurred (Fig 1). From 1960-1992 there were 1511 cases and 478 deaths (CFR 31.6%) that occurred in 11 of Ghana’s 16 regions. The greatest number of YF cases were reported in Upper West (n=384), Volta (n=340), Eastern (n=250), Upper East (n=163), Savannah (n=124), and Bono (n=105) regions. Following implementation of routine childhood YF vaccination in 1992, the mean annual number of YF cases fell by 81%. From 1992-2022 there were 291 cases and 70 deaths (CFR 24.1%) with the highest number of cases in the Upper West (n = 135), Savannah (n = 45), and Upper East (n=33) regions and few cases in the southern regions. The districts where YF outbreaks were suspected to originate in Ghana based on primary/index cases are shown in Fig 2.

**Fig 1.**
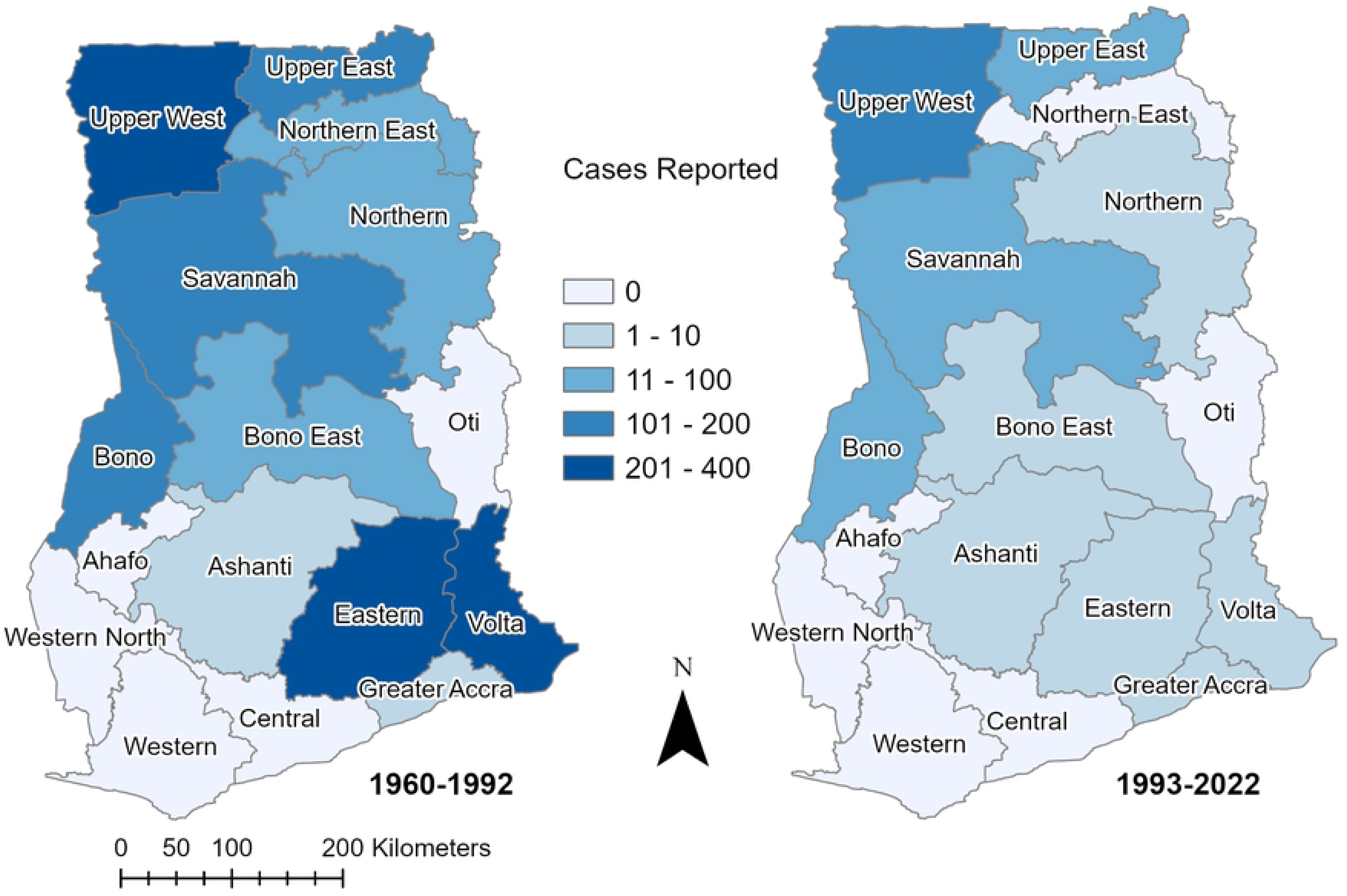
Yellow Fever Cases Reported by Region in Ghana. Reported YF cases that could be located on a regional level in Ghana are shown for the periods before and after routine childhood YF vaccination, 1960-1992 and 1993-2022. The regions of historical cases have been updated to match the current terminology for the 16 regions in Ghana.

**Fig 2.**
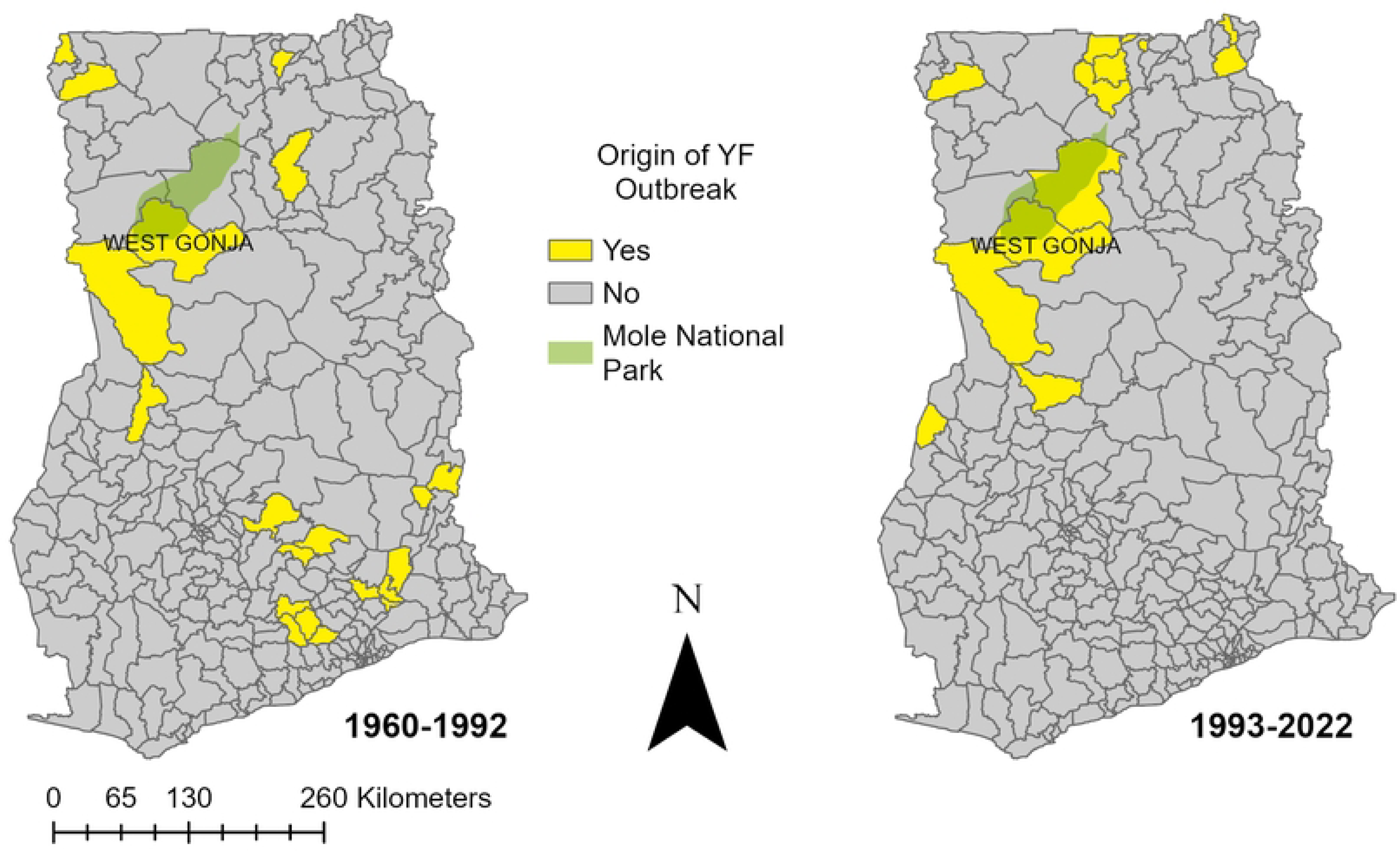
Yellow Fever Outbreak Origins by District in Ghana. The districts where reported YF outbreaks were suspected to originate in Ghana based on primary/index cases are shown for the periods before and after routine childhood YF vaccination, 1960-1992 and 1993-2022. Also shown is Mole National Park and the West Gonja District where the 2021 and 1983 outbreaks were suspected to originate. For the purposes of this figure, outbreaks are defined as more than one case of YF.

We identified certain outbreaks and cases that could be categorized as likely due to urban, savanna, and sylvatic cycles in Ghana (Table 2). Of eleven outbreaks with available epidemiological data that occurred between 1910 and 1979, we estimated that nine likely corresponded to the urban cycle. One outbreak (1983) had evidence of starting in the sylvatic/savanna cycles and then spreading through the urban cycle, and at least two recent outbreaks appear to have originated via the sylvatic/savanna cycles. In addition to these outbreaks there have been sporadic individual cases likely from the sylvatic/savanna cycles.

**Table 2.**
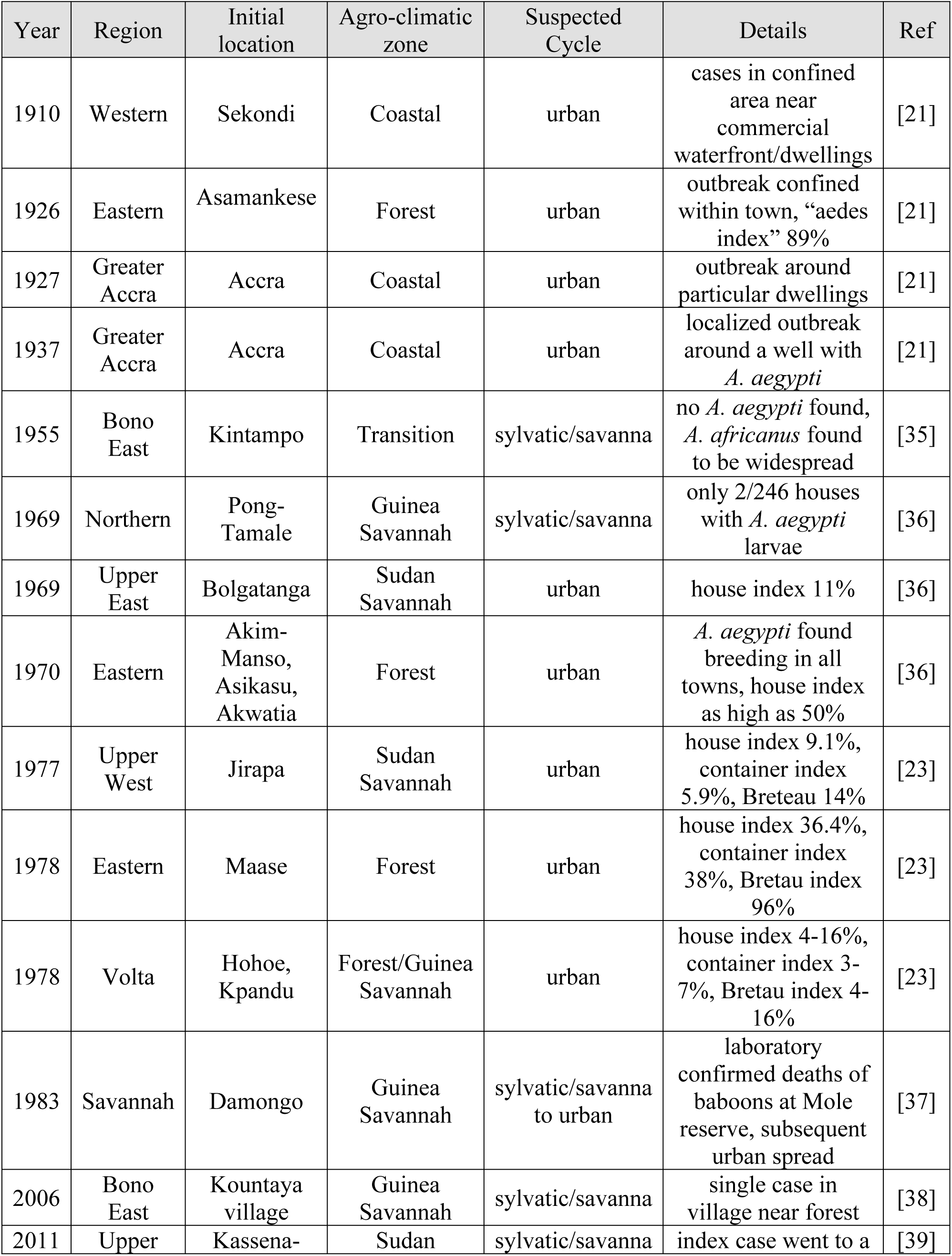
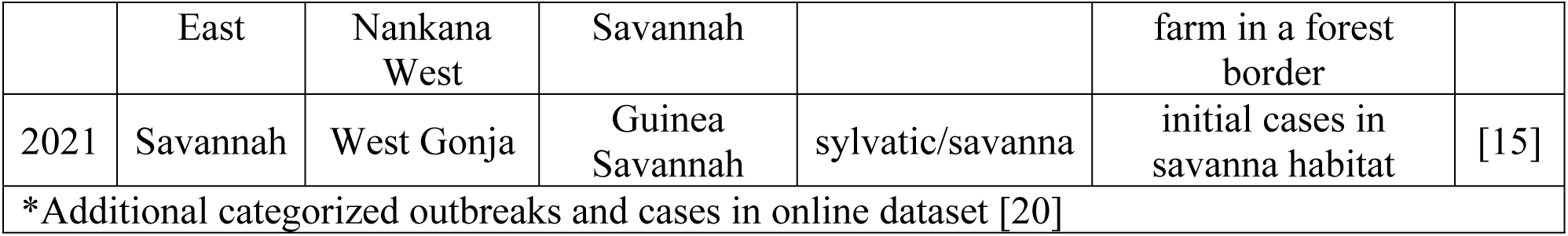
Select YF outbreaks and cases in Ghana with epidemiologic details from 1910-2022*.

The covariates selected for the final models and their relative contributions are shown in Table 3. The average habitat suitability models for YF in Ghana are shown in Fig 3. The minimum training presence threshold was used to translate these models into the binary maps in Fig 4. The overall YF model (AUC 0.846) included landcover, precipitation of the driest quarter (BIO17), human population density, annual mean temperature (BIO1), and elevation. The model that excluded confirmed urban YF occurrences (AUC 0.880) also included landcover and precipitation of driest quarter (BIO17), as well as precipitation of wettest quarter (BIO16) and mean temperature of the driest quarter (BIO9). The most important type of landcover based on the response curves for the models was dense herbaceous vegetation. Human population density had the highest permutation importance and jackknife variable importance for the overall YF model. Precipitation of the driest quarter (BIO17) had the highest permutation importance and jackknife variable importance for the model excluding urban YF. The overall YF model predicted 20/21 (95%) of occurrences tested at the minimum training presence threshold. The model excluding urban YF predicted 14/15 (93%) of occurrences at the same threshold. The only location that was not predicted by both models at the minimum training presence threshold was a single case of likely sylvatic YF in Sene district of the Bono East region. Both models identified regions and districts with recent YF outbreaks as having high suitability including Upper West, Upper East, North East, Savannah, Northern, Bono, and Bono East regions. The overall YF model identified all regions in Ghana as having areas of suitable habitat for YF.

**Fig 3.**
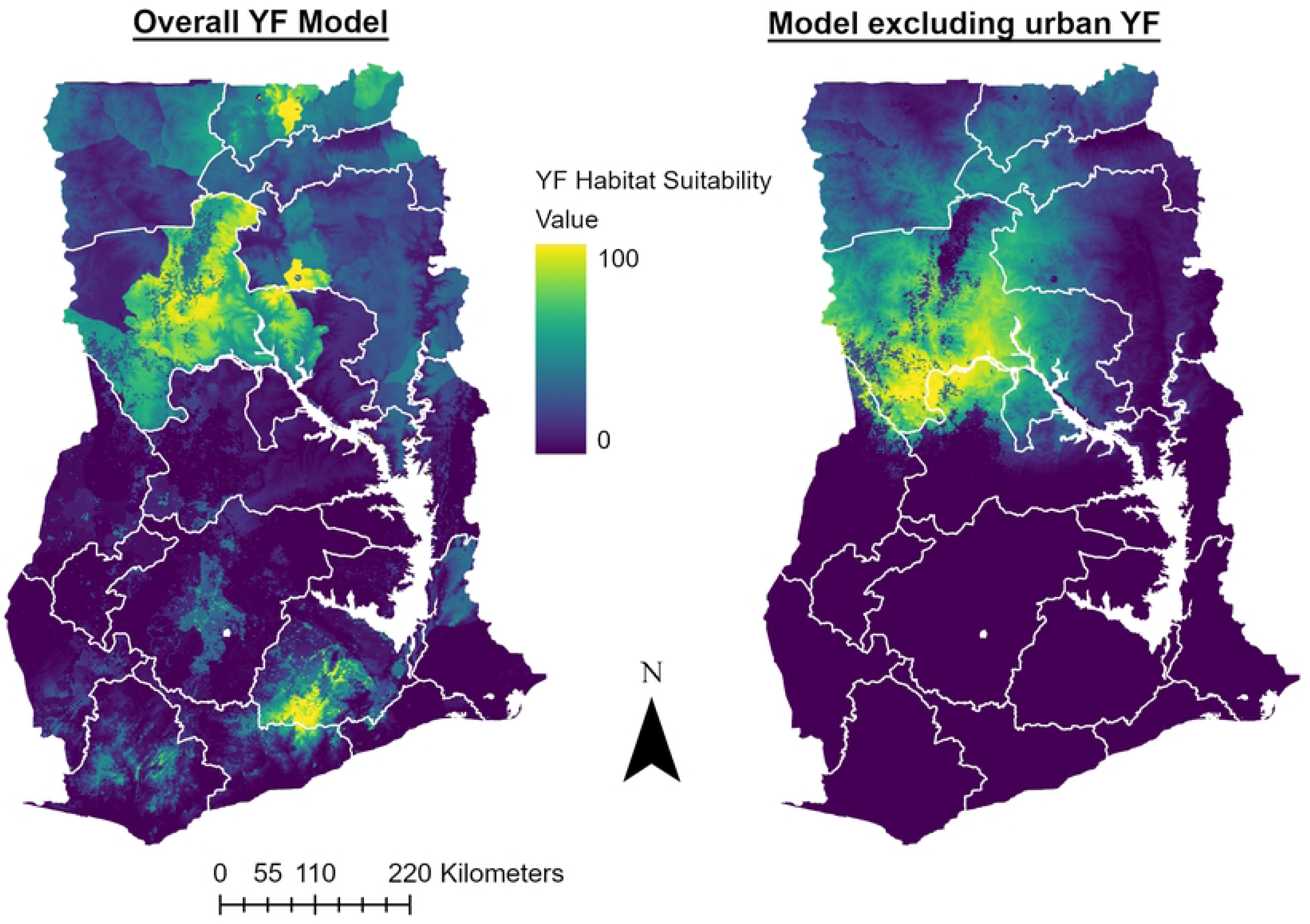
Predicted Habitat Suitability for YF in Ghana. The left panel shows the average Maxent cumulative output for models including 21 YF occurrences in Ghana and the following covariates: human population density, elevation, landcover, precipitation of driest quarter (BIO17), and annual mean temperature (BIO 1). The right panel shows the average output of the models excluding 6 confirmed urban YF occurrences and includes the following covariates: precipitation of driest quarter (BIO17), precipitation of wettest quarter (BIO16), landcover, and mean temperature of driest quarter (BIO9).

**Fig 4.**
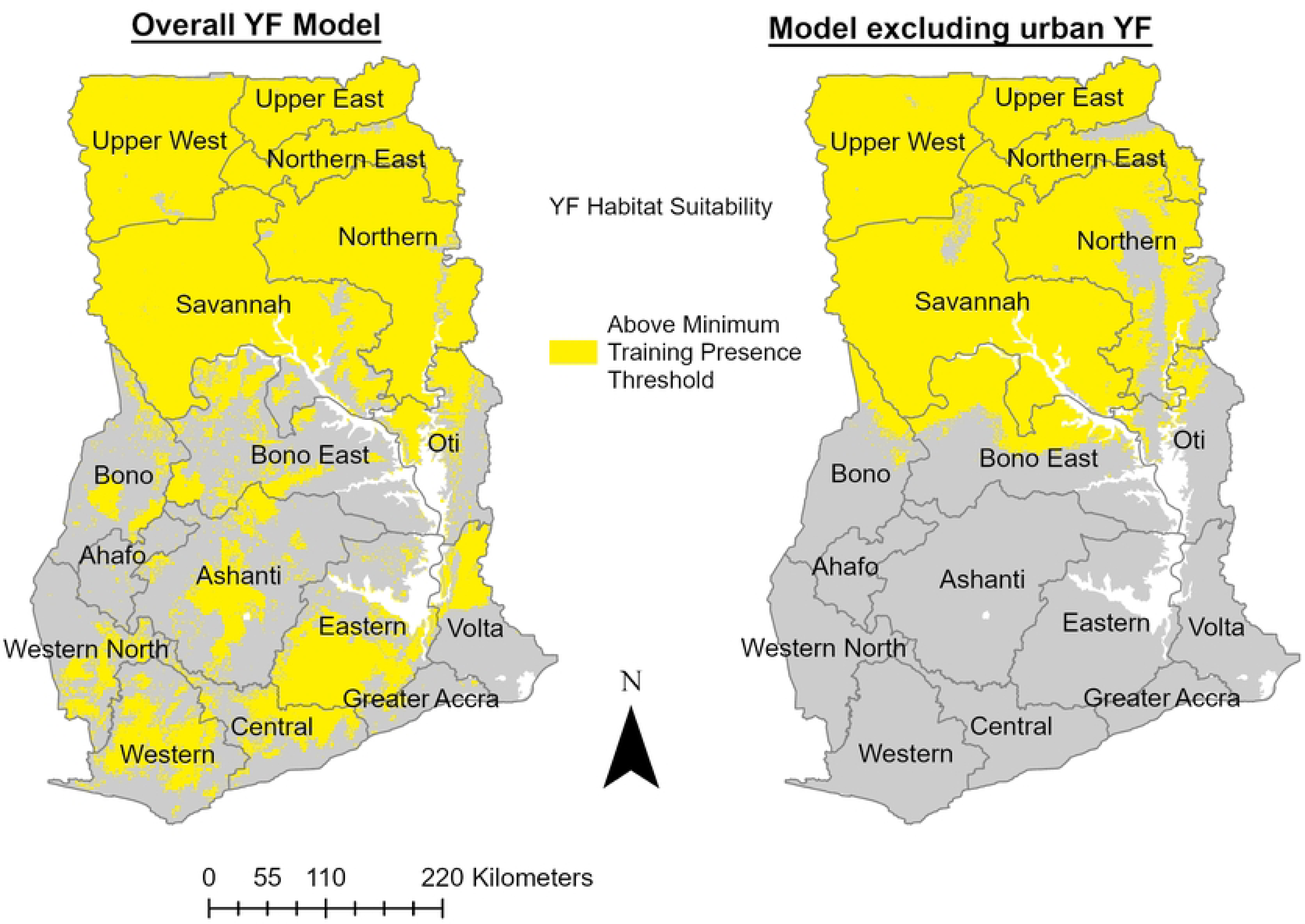
Predicted Habitat Suitability for YF in Ghana based on the Minimum Training Presence Threshold. These binary habitat suitability maps represent areas greater or equal to the minimum training presence threshold (MTP) for each group of YF models. The MTP is defined as the lowest predicted habitat suitability value for the occurrences in the training data. Therefore, the highlighted areas above the MTP represent locations with ecological conditions at least as favorable as the location with the least suitable known YF occurrence in Ghana.

**Table 3.**
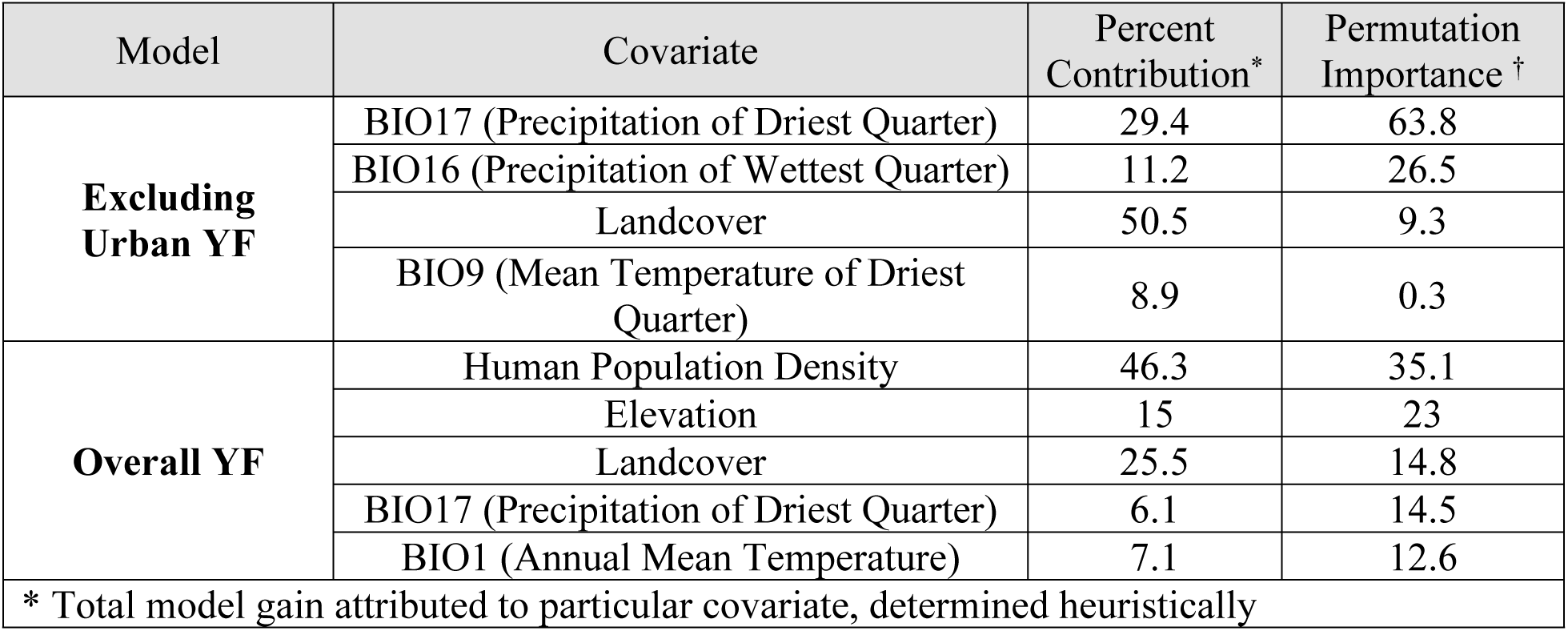
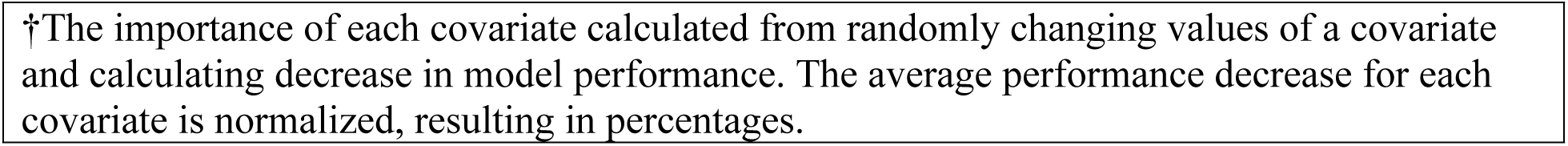
Contribution of final covariates to YF habitat suitability models.

| Model | Covariate | Percent Contribution* | Permutation Importance † |
| --- | --- | --- | --- |
| **Excluding Urban YF** | BIO17 (Precipitation of Driest Quarter) | 29.4 | 63.8 |
|  | BIO16 (Precipitation of Wettest Quarter) | 11.2 | 26.5 |
|  | Landcover | 50.5 | 9.3 |
|  | BIO9 (Mean Temperature of Driest Quarter) | 8.9 | 0.3 |
| **Overall YF** | Human Population Density | 46.3 | 35.1 |
|  | Elevation | 15 | 23 |
|  | Landcover | 25.5 | 14.8 |
|  | BIO17 (Precipitation of Driest Quarter) | 6.1 | 14.5 |
|  | BIO1 (Annual Mean Temperature) | 7.1 | 12.6 |

\* Total model gain attributed to particular covariate, determined heuristically
†The importance of each covariate calculated from randomly changing values of a covariate and calculating decrease in model performance. The average performance decrease for each covariate is normalized, resulting in percentages.

## Discussion

Our analysis estimates the historical epidemiology and ecology of YF in Ghana as well as areas at potential risk for future emergence of YF. We found that the geographic distribution and disease burden of YF in Ghana have changed since the introduction of routine childhood vaccination in 1992. While many historical outbreaks have occurred through the urban cycle, recent outbreaks appear to have originated from the sylvatic or savanna cycles. Therefore, there appears to be a higher risk for YF outbreaks originating among unvaccinated populations in environments where the appropriate vectors, NHPs, and human hosts are present to maintain the sylvatic or savanna cycles. Using machine learning we identified these potential suitable habitats on a more granular scale in Ghana as well as factors that should be considered when predicting YF based on different transmission cycles.

The overall mean CFR we calculated for YF in Ghana (37.4%) was similar to the mean CFR from a recent meta-analysis of global YF cases (39%) and severe cases in African countries (36%) [5]. Given that there is limited surveillance and diagnostic capacity for YF in Ghana, our data likely underestimate of the true burden of YF (while overestimating the true CFR) given that mild or subclinical infections may not be detected. The YF cases in our study more likely reflect severe YF, and it is estimated that ∼15% of YF cases are from severe YF. If only severe YF cases were captured in our dataset, this would suggest that over 15,000 cases of YF have occurred in Ghana since 1910 – a number that may be even higher given historical and current limitations in diagnosing YF. Both the number of cases and the case-fatality of YF have declined in Ghana since 1992, likely in part due to introduction of routine childhood YF vaccination in 1992. Nevertheless, YF outbreaks continue to occur, highlighting the need for ongoing risk prediction and surveillance.

The geographic distribution, epidemiology, and ecology of YF outbreaks in Ghana has changed substantially over the past century. During the first half of the 20^th^ century, the majority of YF outbreaks were detected along coastal regions and periodically in the north, with large outbreaks occurring roughly every 10 years [21]. Relatively higher population density and the presence of susceptible foreign populations likely contributed to these urban YF outbreaks.

Detection bias also likely contributed to these observations given differences in accessibility to diagnostics and reporting. The requirement for YF immunization among foreigners in 1945, local reactive vaccination campaigns, and increasing diagnostic recognition likely contributed to the changing distribution of YF outbreaks with greater recognition of outbreaks in more remote northern regions. In the context of increasing vaccination coverage for YF in Ghana, recent outbreaks have originated among unvaccinated populations in rural areas and border districts in the Guinea Savannah and Sudan Savannah agro-climatic zones. These areas may have more nomadic pastoralist communities and as well as immigrants from neighboring countries, potentially contributing to lower vaccination coverage. Recent YF outbreaks appear to reflect the sylvatic and savanna cycles, which could transition into the urban cycle if there are sufficient susceptible hosts. We found at least one large historical outbreak that began in the sylvatic/savanna cycle and then spread to multiple other regions through the urban cycle. Climate change, urbanization, and changing human population movements could fuel the spread of sylvatic/savanna YF to urban outbreaks.

The Upper West, Savannah, and Upper East regions have experienced the majority of YF cases in outbreaks since routine immunization. Our models identified these regions as well as neighboring regions (Upper East, North East, Northern, Bono, and Bono East) as being highly suitable for YF emergence. The West Gonja district where the 2021 YF outbreak originated is located near Mole National Park (Figure 2), which is the largest protected area in Ghana and is likely suitable habitat for the NHPs and mosquitoes that could maintain the sylvatic and savanna cycles of YF. Mole National Park is also a tourist site where nomadic populations may work and also has surrounding pastoralist communities [40]. The 1983 YF outbreak was also believed to have originated from this area given the finding of laboratory confirmed deaths of baboons with YF [37]. Reviewing the locations of historical YF emergence, we found additional locations with recurring cases, which could also be hotspots for YF. At least four outbreaks appear to have originated in the Jirapa district in the Upper West and three in Bawku Municipal district in the Upper East. During the 2021 outbreak there were two cases from Tinga village in the Bole region, and given the distance from the epicenter of the outbreak it is possible that this could have been a separate YF emergence event. This village also had clinically diagnosed cases of YF in 1983. While there have been isolated cases of sylvatic/savanna YF since routine immunization, with index cases exposed to forest-savanna ecotones, these cases did not lead to sustained outbreaks. In contrast, a significant unvaccinated population likely contributed to the 2021-2022 outbreak. Therefore, identifying both susceptible populations as well as the habitat for YF emergence will be crucial for predicting areas at future risk.

We used confirmed human cases and machine learning to identify likely suitable habitats for the emergence of YF in Ghana. The models do not represent the distribution or ecological niche of YFV, but instead represent habitats with similar conditions to where prior YF cases occurred. Given that YF can start in the sylvatic/savanna cycles and then spread through urban cycle, we also created a set of models excluding confirmed urban YF cases to provide a better prediction of habitats where outbreaks may originate. This model is a proxy for the habitats similar to where savanna/sylvatic cases have occurred, which appears to be the likely origin of recent and future YF outbreaks in Ghana. Because we used the location of residence or healthcare facility for our occurrences, the habitats identified by this model do not reflect where YFV circulates in the savanna/sylvatic cycle, but instead habitats similar to where human cases from these cycles have been detected.

To our knowledge, this is the first time that the different ecological cycles of YF have been integrated into spatial models in Africa. Overall these models illustrate the important differences in habitats and locations where YF may occur in Ghana. For example, while landcover, precipitation, and temperature are important covariates in both sets of models, human population density was the most important covariate for models that included outbreaks from the urban cycle– reflecting the fact that the urban cycle requires a higher density of susceptible human hosts compared to the sylvatic/savanna cycles. In contrast, precipitation of the driest quarter was the most important covariate for the model that excluded urban YF. Predictive models that do not distinguish between the ecological cycles of YF will risk misidentifying areas at risk for emergence. For instance, our models excluding urban cases reflect habitat similar to where the recent YF outbreaks have originated in Ghana and could be a better predictor for where future outbreaks may originate. In contrast, the overall model is more appropriate for predicting areas where YF may spread through the urban cycle. Therefore, it is important that researchers think carefully about the goals and inferences of their models based on YF ecology and epidemiology.

There are multiple limitations to our analysis. We used reported YF cases from the literature and online reports, and there were relatively few laboratory confirmed historical cases. For our habitat suitability models, we used strict inclusion criteria of confirmed cases since 1960 with georeferenced locations, limiting our analysis to 21 total occurrences. We used AUC values to compare the relative performance between models, but AUC values can be inflated in presence-only models such as Maxent. We attempted to minimize this inflation and reduce overfitting by using the LOOCV approach and a small number of covariates. Another limitation was sparse epidemiologic and entomologic surveillance data for most outbreaks. Vector indices were only available for some of the occurrences, so we had to make assumptions about certain outbreaks that lacked entomologic data, and we only excluded confirmed urban YF locations in our second set of models. Similarly, it is possible that both urban and savanna/sylvatic cycles may occur together during outbreaks. For example, while the 2021 YF outbreak may have originated in the savanna/sylvatic cycle, it could have spread through dwellings via the urban cycle. We categorized this outbreak and similar outbreaks as originating from the savanna/sylvatic cycle since our goal was to model habitat for YF emergence. The alternative explanation, that this YF outbreak originated from the urban cycle of another outbreak, seemed less likely given lack of nearby preceding outbreaks. Overall, additional vector surveillance data are needed for further clarity. Lastly, the covariates we chose for our models were intended to reflect habitat suitability via presence of the appropriate climatic conditions, environments, and hosts. To make models more reflective of relative risk for future outbreaks, they would need to include additional explanatory variables, most importantly YF vaccination coverage.

Despite these limitations, our study has important implications for preparing for future outbreaks of YF in Ghana. For example, our models could be used to inform further risk assessment which could guide diagnostic testing, vector control, and vaccination campaigns. A committee of YF experts developed a WHO protocol for national risk assessment for YF [24]. However, the strategies in this protocol are resource intensive and involve sampling humans, vectors, and NHPs for YF. Our methods for estimating habitat suitability based on historical cases could be used to prioritize field surveillance for YF with the WHO risk assessment protocol [24]. Our findings also have important implications for models and risk maps for YF. While predictive models for YF risk have been made on a global scale [41–43], these models either contain sparse data from Ghana or do not predict subnational risk for YF. Additionally, these existing models do not distinguish between the different ecological cycles of YF in Africa, which may influence prediction and interpretation. Therefore, additional subnational data and local ecological knowledge are needed to create future risk maps for YF and other arboviruses if they are to be informative for national policymakers [9].

While the habitats suitable for YF emergence appear to cluster in the northern regions of Ghana, currently all YF diagnostic testing is done at the National Public Health Reference Laboratory in Korle Bu in the south of the country. Expanding YF diagnostic testing to the Zonal Public Health Laboratory in Tamale, which is close to recent YF outbreaks and predicted habitat suitability, could improve early detection of outbreaks. While the burden of YF has decreased since implementation of routine childhood vaccination in 1992, the recent outbreak has also demonstrated the risk for future outbreaks among unvaccinated groups. Identifying unvaccinated nomadic populations in habitats that are suitable for YF emergence will be an important step for preventing future outbreaks. Overall, despite a century of research on YF, critical questions remain regarding the ecology, epidemiology, and emergence of YF in African countries. Addressing these research gaps and using this knowledge to inform public health interventions will be essential to improving health equity and preventing future YF epidemics.

## Data Availability

All relevant data are available within the manuscript and its online files. Datasets are available and can be cited at the online repository https://doi.org/10.6084/m9.figshare.24747165

## Acknowledgements

The authors would like to thank Dr. Eric Agboli for sharing the 2005 WHO YF in Ghana report and Dr. Sarah Louise Poynton for editing advice. They would also like to thank their colleagues at the Ghana Health Service, University of Ghana, and Noguchi Memorial Institute for Medical Research.

## Author contributions

Conceptualization, Data curation, Analysis, Writing-original Draft: SDJ

Methodology: SDJ, TF

Writing-reviewing and editing: SDJ, DWD, LFS, TF, FA, EK, AB

Funding: DWD, LFS, FA, EK

## Supporting Information

**S1 Fig. Yellow Fever Annual Cases and Deaths in Ghana 1910-2022**

The reported annual number of YF cases and deaths in Ghana since the first detected outbreak in 1910 until 2022 are shown. Also depicted are the years of reactive YF vaccination campaigns and immunization policies.

